# Carotid perivascular fat attenuation on CT angiography as an imaging biomarker for inflammation in carotid atherosclerosis: a systematic review and meta-analysis

**DOI:** 10.1101/2025.10.16.25338115

**Authors:** Claudia Zeicu, Shiv Bhakta, Jason M Tarkin, Jacob Brubert, James HF Rudd, Elizabeth A Warburton, Nicholas R Evans

## Abstract

**Aims:** Carotid atherosclerosis is a leading cause of ischemic stroke, driven by plaque inflammation. While current carotid imaging focuses on luminal stenosis, quantitative perivascular adipose tissue attenuation (PVAT) on computed tomography angiography has emerged as a potential non-invasive biomarker for arterial inflammation and plaque instability in coronary arteries. This systematic review and meta-analysis consider the utility of PVAT for imaging and risk-stratifying inflamed and vulnerable carotid atherosclerotic plaques.

**Methods and Results:** Following PRISMA guidelines, a systematic literature search identified 11 studies suitable for meta-analysis, incorporating 2316 patients. We compared continuous PVAT measures between symptomatic and asymptomatic carotid arteries using mean differences and 95% confidence intervals, employing a random-effects meta-analysis due to substantial heterogeneity (I² = 92%). The pooled mean difference (MD) was 12.63 HU (95% CI: 8.39-16.88), indicating significantly higher PVAT in symptomatic arteries. While funnel plot asymmetry was visually observed, Egger’s test (intercept = 7.29, p = 0.044) confirmed statistically significant publication bias, highlighting the need to apply caution when interpreting the results given the high heterogeneity. Notably, the leave-one-out sensitivity analysis showed no significant change in the overall p-value or substantial shifts in statistical heterogeneity.

**Conclusion:** This meta-analysis supports PVAT as a promising non-invasive imaging marker for inflammation and vulnerable plaques in symptomatic carotid atherosclerosis. The findings, while subject to heterogeneity and requiring further validation against conflicting individual study results, suggest a possible role of carotid PVAT for risk stratification. Future directions may PVAT use for monitoring treatment response in clinical trials.

## Introduction

Stroke remains a leading cause of death and long-term disability worldwide, accounting for a significant global health burden [1, 2]. A major aetiology for ischemic stroke is carotid atherosclerosis, accounting for approximately 20% of strokes [3], where the rupture of unstable (“vulnerable”) atherosclerotic plaques results in thromboembolic events [4, 5]. Inflammation within the plaque microenvironment is an important driver of this instability, promoting neovascularisation, haemorrhage, and fibrous cap thinning which increase the risk of plaque rupture and stroke, and is a strong predictor of recurrent stroke [6–9]. Consequently, identification and timely treatment of vulnerable atherosclerotic plaques is critical for stroke risk reduction.

Conventional imaging methods used to evaluate the carotid arteries in clinical practice, such as ultrasound and computed tomography angiography (CTA), primarily quantify luminal stenosis, a parameter that determines surgical intervention but does not necessarily reflect the underlying inflammatory activity or plaque vulnerability [10, 11]. Consequently, high-risk inflamed plaques with sub-threshold stenosis may be overlooked and/or the risk over-estimated from stable but highly stenosed plaques using a solely stenosis-based paradigm [12]. Imaging techniques that can provide evaluation of high-risk pathophysiological or morphological plaque features are highly desirable, but techniques used in research (such as positron-emission tomography and high field-strength magnetic resonance imaging) have failed to make the translational leap to clinical practice due to a combination of cost, radiation exposure, and availability.

Assessing perivascular fat attenuation using CT is a technique used in coronary artery imaging that may have a use in carotid imaging. Coronary studies indicate that inflamed atheroma may alter the attenuation signal intensity in adjacent perivascular fat. [13]. These signal changes are caused by increased attenuation values on CTA due to local oedema and lipolysis [14]. The concept of a perivascular fat attenuation index has shown both diagnostic and prognostic applications in coronary artery disease [15]; however, its use in the carotid arteries is less clear, and evidence is limited to a small number of studies. Given the increasing use of CTA in acute imaging protocols in stroke – such as the National Optimal Stroke Imaging Pathway (NOSIP) in the United Kingdom – a CTA-based technique to identify perivascular fat attenuation using imaging acquired in routine clinical practice is highly desirable and more straightforward to implement clinically than an additional dedicated imaging test to assess inflammation.

Considering the limitations of current stroke risk-stratification tools, and the emerging evidence regarding the role of perivascular fat attenuation index in carotid atherosclerosis, we conducted a systematic review and meta-analysis to clarify the potential of non-invasive imaging markers in the identification of carotid plaque instability, and provide further proof-of-principle for the technique in improving the evaluation of stroke risk from carotid atherosclerosis.

## Materials and Methods

### Search strategy and selection criteria

The systematic review was designed according to the Preferred Reporting Items for Systematic Review and Meta-Analysis (PRISMA) checklist [16]. A comprehensive search was undertaken using the search terms detailed in Table 1, covering studies published from inception up to June 1st, 2025. The objective of the review was to understand if the adipose or fat tissue attenuation noted on CT angiography of patients admitted with ischemic strokes was related to symptomatic carotid atherosclerosis.

**Table 1.**
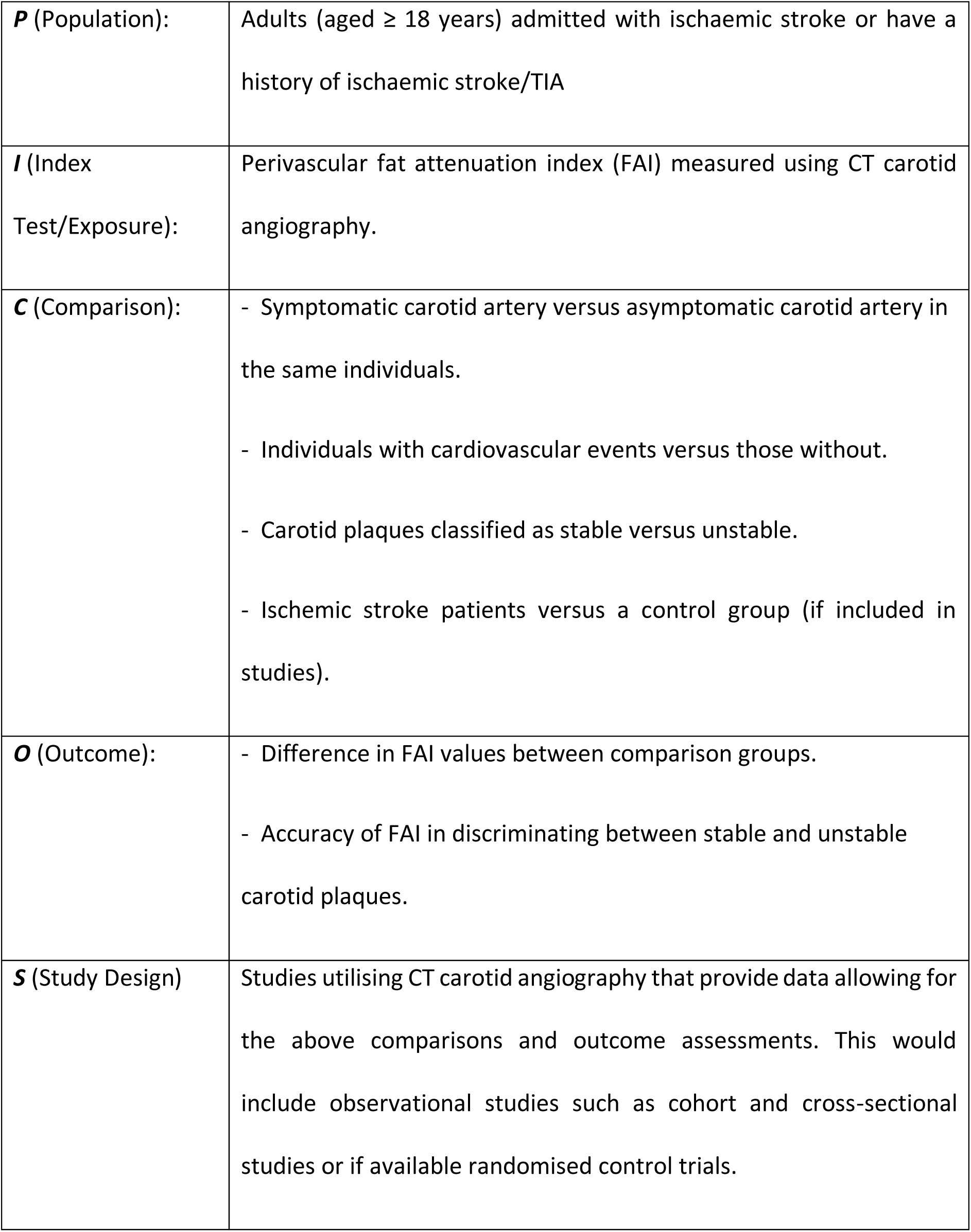
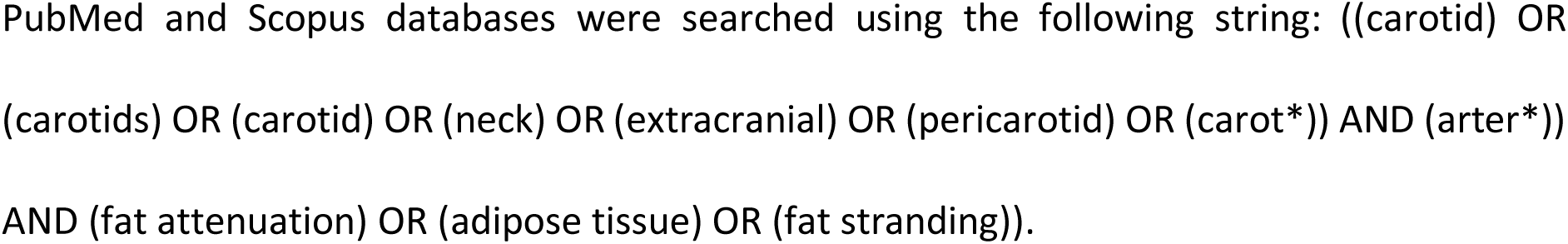
PICOS inclusion criteria.

Studies were included if they involved adults (aged ≥18 years) who underwent CT carotid angiography and had a measured perivascular fat density index, and who were either admitted with acute ischemic stroke or had a history of ischemic stroke or transient ischemic attack (TIA). To ensure focus on carotid atherosclerosis, studies were excluded if they did not explicitly compare the symptomatic versus contralateral asymptomatic carotid artery or a control group, or if the stroke aetiology on the symptomatic side was determined to be other than large vessel atherosclerosis. Studies using positron emission tomography (PET) instead of CT angiography were also excluded. Case reports or case series with fewer than 10 cases, preprints, conference abstracts, communications, letters to the editor, and reviews were not eligible for inclusion.

### Data extraction

Selected studies included manuscripts published in peer-reviewed journals, with an abstract available, without language limitations. Search results were exported into Excel (Office Suite, Microsoft Corp, Redmond, Washington, USA). After duplicates were removed, two independent reviewers screened titles, abstracts, and full-text articles for eligibility criteria. All reference lists from included articles were screened to identify additional pertinent studies. For the included papers, data regarding patients’ demographics, exclusion criteria, and relevant findings were extracted. The selection process was outlined in a PRISMA flow diagram (Figure 1). Funnel plot and Egger regression test were used to assess publication bias.

**Figure 1.**
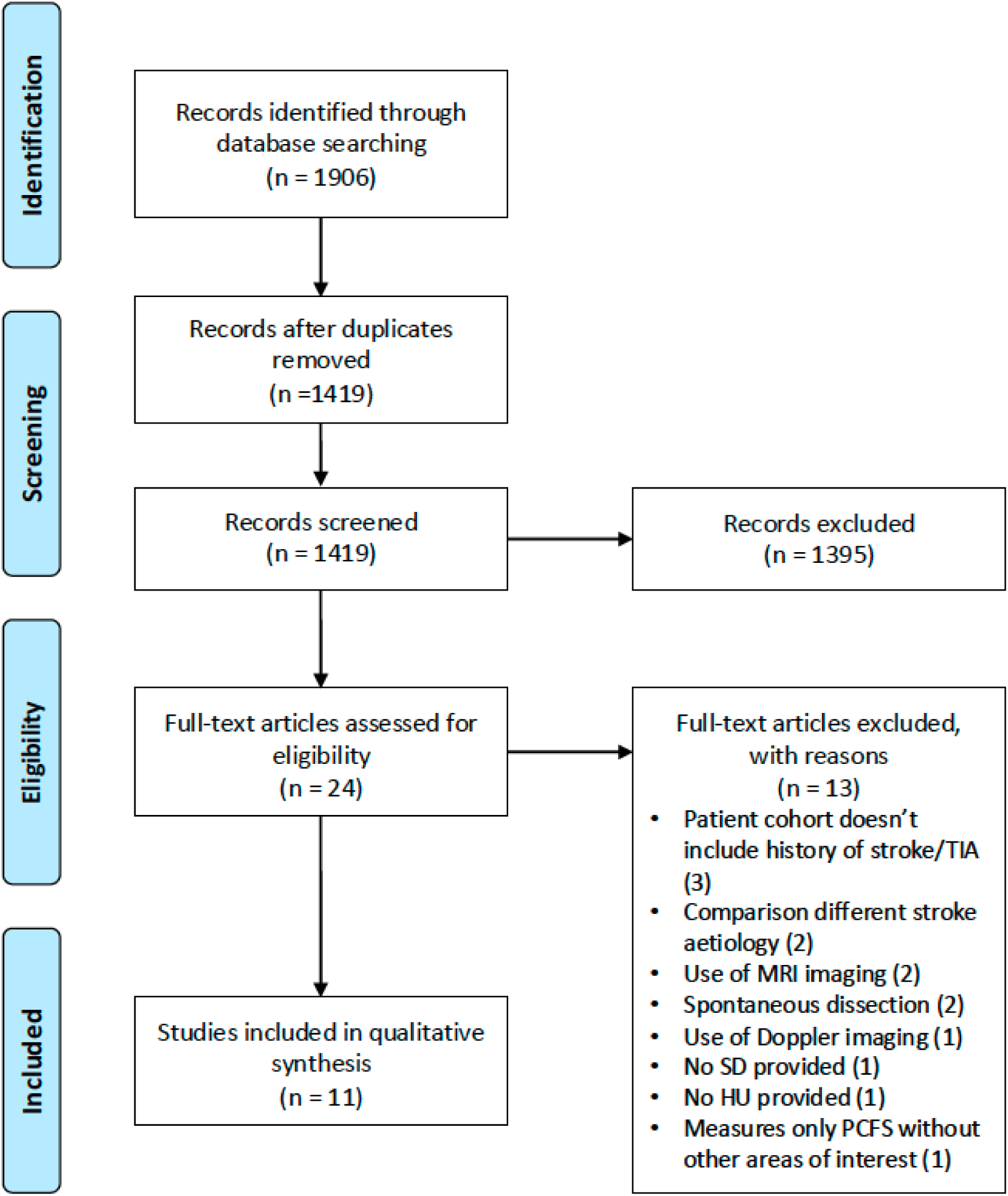
PRISMA flow diagram illustrating the study selection process for the meta-analysis. TIA - transient ischaemic attack, MRI – magnetic resonance imaging, SD – standard deviation, HU-Hounsfield units, PCFS – pericarotid fat stranding.

The review protocol was registered in the PROSPERO registry, reference number CRD420250653431.

### Statistical analysis

Continuous perivascular fat attenuation (PVAT) measures were compared between symptomatic and asymptomatic carotid arteries using the mean difference (MD). For each study, the MD, and 95% confidence interval (CI) were calculated, with standard errors (SE) derived from the reported standard deviations (SDs). One study [17] reported their data as medians and interquartile ranges, necessitating an estimation methodology to derive the mean and standard deviation for meta-analytic inclusion [18].

Statistical heterogeneity across studies was assessed using Cochran’s Q and the I2 statistic. Both fixed-effects (inverse variance) and random-effects meta-analyses were conducted to estimate the pooled MD. The random-effects model specifically utilised the simplified DerSimonian-Laird method, incorporating inverse-variance weighting. Given the degree of heterogeneity (I2 <25% for low, ≥25% for moderate to high), the random-effects pooled MD was chosen for primary interpretation. The pooled MD and heterogeneity statistics were calculated using Python (version 3.10.9) within the Spyder Integrated Development Environment (version 5.15.2), utilising the pandas, numpy, scipy.stats, and statsmodels libraries. The forest and funnel plots were created in Spyder IDE using matplotlib and forest plot libraries, and the subsequent Egger test utilised the statsmodels library. The meta-analysis reliability and validity were assessed using leave-one-out sensitivity analysis.

## Results

The study selection process is illustrated in the PRISMA flow diagram (**Figure 1**). Initial literature searches identified 1906 studies, which contained 487 duplicates, with 24 studies proceeding for full-text review. The systematic review and meta-analysis included 11 studies, incorporating data from a total of 2316 patients.

The key characteristics of these studies, including author, year of publication, number of subjects, the proportion of male participants, mean age of participants, exclusion criteria, and relevant findings, are summarised in Table 2.

**Table 2.** Baseline characteristics of the included studies (SD – standard deviation, PVAT – perivascular adipose tissue attenuation, HU – Hounsfield units, FAI – fat attenuation index).

| Author<br>(year) | Subjects | Male (%) | Mean<br>Age, SD | Exclusion<br>Criteria | Relevant Findings | PVAT method | Software | Absolute HU or<br>FAI |
| --- | --- | --- | --- | --- | --- | --- | --- | --- |
| Baradaran et al.<br>(2018)<br>[21] | 94 patients, 42 symptomatic carotid plaques | 58 (61.7%) | 73.4 ± 9.9 | (1) any patients who had simultaneous bilateral anterior circulation events (2) patients with bilateral extracranial ICA stenosis. | Symptomatic subjects had significantly higher mean pericarotid fat density (−66.2±19.2 vs. −77.1±20.4 HU, p=0.009) | Two 2.5 mm<sup>2</sup> regions of interest (ROIs). | Not explicitly stated. | Absolute HU values. |
| Saba *et al.* (2020) [22] | 100 patients, 31 symptomatic | 76 (76%) | 69 | (1) subjects younger than 18 years of age; (2) CTAs performed for reasons other than suspected atherosclerotic disease (i.e. dissection); (3) other aetiologies for ischemic stroke such as evidence of a cardiac embolic source, evidence of an embolism from the thoracic aorta, and evidence of vertebrobasilar artery disease. | Symptomatic subjects had higher mean perivascular fat density (PFD) (-67.1±8.0 vs -77.9±5.9). | Two regions of interest, each 2.5 mm<sup>2</sup>. | Not explicitly stated. | Absolute HU values. |
| Zhang<br>*et al.*<br>(a) (20 | 72<br>patients<br>with 47 | 56<br>(77.78%) | 65.1 ±<br>9.1 | (1) diseases other than atherosclerotic disease (i.e. dissection); (2) | Compared with carotid plaques without symptoms, those with symptoms had higher PFD (−33.5 | Semi-automatically tracked with | Perivascular Fat Analysis Tool (Shukun | Absolute HU values. |
| 21)<br>[23] | symptom<br>atic | | | history of<br>carotid<br>endarterectomy<br>and stenting;<br>and (3) CTA and<br>magnetic<br>resonance (MR)<br>images with<br>poor quality. | $\pm 11.6$ vs. $-54.1 \pm 8.9$<br>HU; $p < 0.001$). | dedicated<br>software. | Technology,<br>Beijing, China). | |
| Zhang<br>*et al.*<br>(b)<br>(2021)<br>[24] | 572<br>patients,<br>377 with<br>acute | 373<br>(65.2%) | 66 | (1) severe<br>malacia lesion,<br>extensive or old<br>infarction,<br>haemorrhage,<br>atrophy or | Subjects with acute<br>stroke had a higher<br>mean pericarotid fat<br>density than without<br>($-76.1 \pm 13.0$ vs. - | Two regions<br>of interest, 3<br>$\text{mm}^2$ in<br>diameter. | Not explicitly<br>stated. | Absolute HU<br>values. |
|  | ischemic<br>stroke |  |  | tumour; (2)<br>other diseases<br>that may cause<br>white matter<br>lesions, such as<br>multiple<br>sclerosis,<br>vasculitis, and<br>connective<br>tissue diseases;<br>(3) poor imaging<br>quality or<br>partially missing<br>images. | 79.6±12.8 HU,<br>p=0.002). |  |  |  |
| Hu *et al.* (2022) [25] | 244 patients, 126 with embolic stroke of undetermined source (ESUS), 118 large artery atherosclerosis (LAA) | ESUS (65.87%)<br>LAA (80.51%) | ESUS: 68.11 ± 10.33,<br>LAA: 68.82 ± 11.20 | (1) simultaneous bilateral anterior circulation events; (2) patients with simultaneous anterior and posterior circulation strokes; (3) patients with intracranial large artery stenosis; (4) | Higher perivascular fat density around the carotid artery ipsilateral to the stroke vs. the contralateral in both the groups (ESUS cases -56.3 ± 18.7 vs. -67.3 ± 20.0, p=0.000; LAA ischemic stroke cases -51.6 ± 20.0 vs. -64.6 ± 22.7, p=0.000) | Two regions of interest, each 2.5 mm<sup>2</sup>. | Not explicitly stated. | Absolute HU values. |
|  |  |  |  | patients without<br>carotid plaque. |  |  |  |  |
| Zhang<br>*et al.* (<br>2022)<br>[26] | 206<br>patients,<br>102<br>symptom<br>atic<br>carotid<br>plaques | 145<br>(70.4%) | 63.9 ±<br>7.9 | (1) history of<br>carotid<br>endarterectomy<br>and stenting, (2)<br>cardiac<br>thrombus, (3)<br>carotid<br>occlusion, (4)<br>posterior<br>circulation<br>symptoms, and<br>(5) CTA images<br>of poor quality; | Subjects with<br>symptomatic plaques<br>have a higher<br>pericarotid fat density<br>than asymptomatic<br>plaques ($-55.0 \pm 10.0$<br>vs. $-68.0 \pm 10.3$ HU,<br>$p < 0.001$) | Semi-automatically<br>tracked with<br>dedicated<br>software. | Perivascular Fat<br>Analysis Tool<br>(Shukun<br>Technology,<br>Beijing, China). | Absolute HU<br>values. |
|  |  |  |  | (6) abnormal lesion at the intracranial arteries on CTA. |  |  |  |  |
| Yu *et al.* (2022) [27] | 71 patients, 42 with acute cerebral infarction | 57 (80.28%) | 61.25 ± 10.35 | (1) patients with diseases other than atherosclerotic disease (e.g., dissection, aneurysm, or Takayasu's arteritis); (2) the CTA and MRI images were<br>poor in quality;<br>(3) patients with a history of carotid endarterectomy and stenting; (4) patients with neoplasms and inflammatory lesions; (5) patients with cardiac, hepatic and/or renal insufficiency. | Symptomatic patients had a higher mean pericarotid fat density compared to asymptomatic ( -36.3 ± 20.7 HU and -66.9 ± 15.0 HU).<br>Intraplaque haemorrhage subgroup had a<br>higher mean HU on the plaque side ($-29.6 \pm 19.2$ vs $-47.7 \pm 18.3$ vs., $p<0.001$). The intraplaque haemorrhage subgroup had a higher prevalence of cerebral infarction group ($p=0.007$). | Two regions of interest, each 2.5 mm². | PACS (GE Healthcare, CENTRICITY PACS 4.0). | Absolute HU values. |
| Liu *et al.*<br>(2023)<br>[28] | 177<br>participants, 45<br>acute<br>stroke<br>patients | 152<br>(85.88%) |  | (1) history of<br>allergy to iodine<br>contrast media<br>or renal<br>dysfunction<br>(estimated<br>glomerular<br>filtration rate<br>assessed by<br>creatinine<br>clearance <60<br>mL/min/1.73<br>m<sup>2</sup>). (2) Aortic<br>artery and<br>ipsilateral anterior<br>circulation<br>intracranial<br>artery<br>arteriosclerosis<br>with $\geq 50\%$<br>stenosis. (3)<br>Evidence of non-<br>atherosclerotic<br>cerebrovascular<br>diseases (e.g.,<br>vasculitis,<br>Moyamoya<br>disease,<br>fibromuscular dysplasia). (4)<br>Echocardiograph<br>y evidence of<br>cardiogenic<br>embolism. (5)<br>History of<br>transluminal<br>intervention. | Subjects with stroke<br>had a higher<br>pericarotid adipose<br>tissue attenuation on<br>the symptomatic side<br>than on the non-<br>symptomatic side<br>(-78.8±11.6 vs. -86.0<br>±11.3 HU, p<0.001) as<br>well as carotid<br>stenosis side of non-<br>stroke patients<br>(-78.8±11.6 HU vs. -<br>89.0±10.8 HU,<br>p<0.001) and no carotid<br>atherosclerotic<br>disease controls<br>( $-78.8 \pm 11.6$ HU vs.<br>$-95.2 \pm 10.8$ HU,<br>$p < 0.001$ ). | Circular<br>regions of<br>interest, each<br>measuring<br>2.5 mm<sup>2</sup>. | VascuCAP<br>software (Elucid<br>Bioimaging,<br>Wenham, MA,<br>USA). | Absolute HU<br>values. |
| Beşler<br>*et al.* (<br>2024)<br>[29] | 374<br>patients,<br>135<br>symptom<br>atic | 234<br>(62.57%) | 68 | (1) purely<br>calcified<br>plaques, where<br>plaque density<br>could not be<br>measured (2)<br>patients who did | Symptomatic subjects<br>had a higher<br>pericarotid fat density<br>compared to<br>asymptomatic group<br>(−63.3 ± 21.7 vs. | A region of<br>interest with<br>an area of 1.3<br>mm<sup>2</sup>. | Advantage<br>Workstation 4.7<br>(GE Healthcare<br>Revolution,<br>USA). | Absolute HU<br>values. |
| | | | | not have<br>plaques<br>detected on<br>carotid CTA | $-81.7 \pm 16.9$ HU,<br>$p < 0.001$). | | | |
| Lan *et al.*<br>(2024)<br>[30] | 134<br>patients,<br>61 had<br>obstructiv<br>e<br>coronary<br>artery<br>disease<br>(CAD) | 94<br>(70.15%) | $60.91 \pm$<br>10.16 | (1) history of<br>carotid<br>endarterectomy<br>or stenting; (2)<br>previous<br>percutaneous<br>coronary<br>intervention<br>(PCI) or<br>coronary bypass surgery; (3) poor image quality | Subjects with<br>obstructive CAD had a<br>mean pericarotid fat<br>density higher than<br>the non-obstructive<br>group ($-52.0 \pm 9.7$<br>vs. $-57.5 \pm 11.3$<br>HU, $p = 0.004$). | Semi-automatically<br>tracked with<br>dedicated<br>software. | Perivascular Fat<br>Analysis Tool<br>(Shukun<br>Technology). | Absolute HU<br>values. |
| Whitti<br>ngton<br>*et al.*<br>(2025)<br>[17] | 48<br>patients,<br>29 culprit<br>plaques<br>vs 50<br>non-culprit<br>plaques | 32 (67%) | 71 ± 11 | (1) impaired renal function (estimated glomerular filtration rate <30 mL/min/1.73 m2), (2) inability to give informed consent or tolerate the scanner protocol, (3) | Internal carotid artery peri-vascular adipose tissue attenuation culprit –63.0 (–71.8, –58.1) vs non-culprit –63.7 (–72.3, –58.2).<br>Values used show median (interquartile range). | Semi-automatically tracked with dedicated software. | Autoplaque version 2.6, Cedars-Sinai Medical Center, Los Angeles, CA, USA. | Absolute HU values. |
|  |  |  |  | women who<br>were pregnant<br>or breast<br>feeding, (4)<br>allergy to<br>iodinated<br>contrast, (5)<br>evidence of<br>haemorrhagic<br>stroke, (6) if<br>participation in<br>the study would<br>result in a delay<br>to carotid endarterectomy<br>surgery |  |  |  |  |

The pooled MD in perivascular fat attenuation (PVAT) measured between the symptomatic and asymptomatic carotid arteries was estimated using a random-effects meta-analysis due to substantial heterogeneity (I^2^ = 92%, Q=125.40, p<0.001). The pooled mean difference was 12.63 HU (95% CI: 8.39, 16.88) (**Figure 2**), indicating that symptomatic carotid arteries exhibit a significantly higher degree of perivascular fat attenuation compared to asymptomatic carotid arteries.

**Figure 2.**
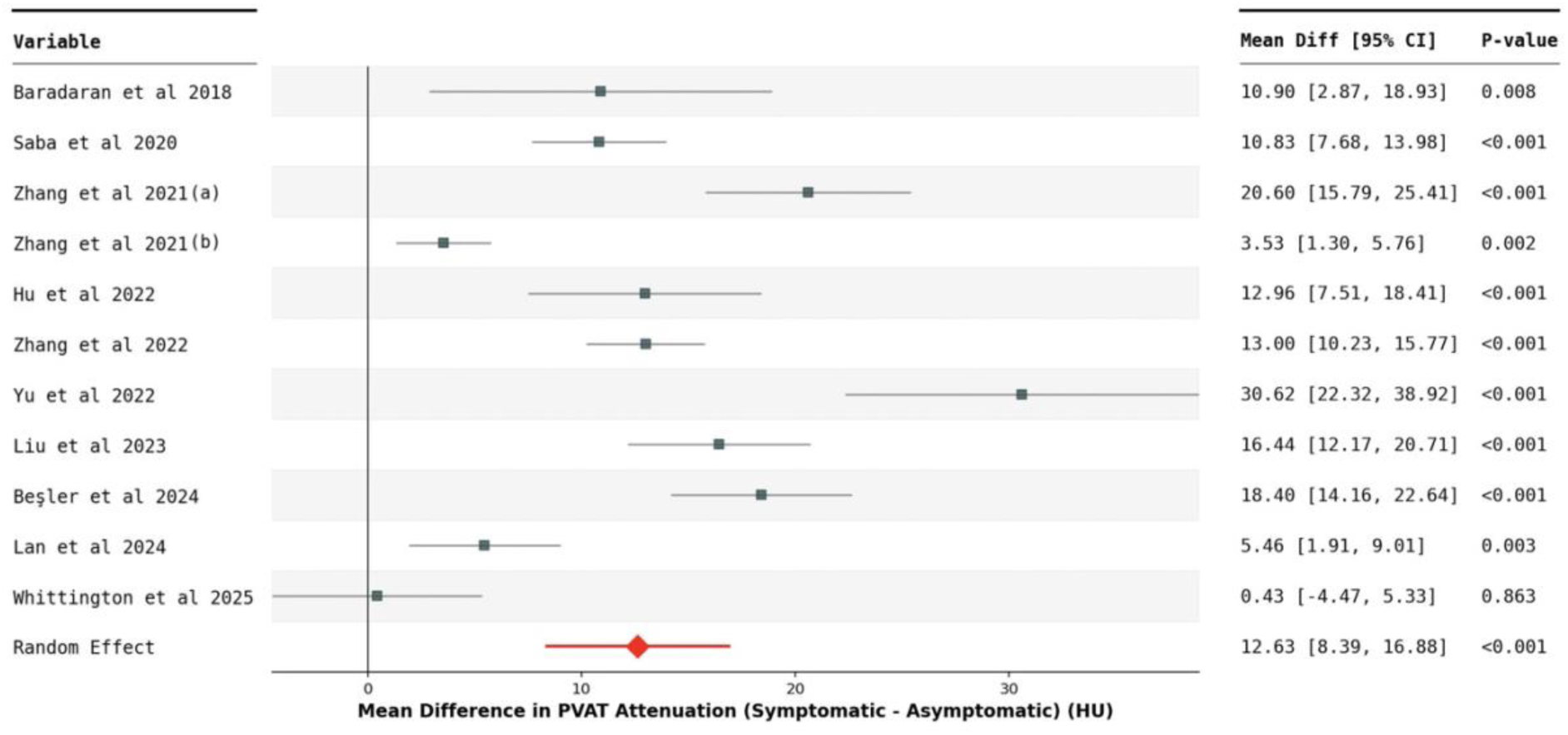
Forest plot showing the mean difference and 95% confidence intervals for individual studies and the pooled mean difference (random effects). The vertical line represents a mean difference of zero (no effect). The red rhomboid shape represents the pooled mean difference from the random-effects model, with its horizontal line indicating the 95% confidence interval. Mean Diff – mean difference, CI – confidence interval.

A funnel plot was generated to assess potential publication bias (**Figure 3**). Visual inspection of the funnel plot revealed some asymmetry, with a potential lack of smaller studies showing negative or non-significant results. Egger’s regression test indicated statistically significant asymmetry (intercept = 7.29, p = 0.0440), suggesting the presence of potential publication bias in the included studies. Given the substantial heterogeneity observed in the meta-analysis (I² = 92%), both the visual assessment and the Egger’s test should be interpreted with caution. To further explore the stability of our findings, a leave-one-out sensitivity analysis was conducted, which demonstrated that the pooled mean difference remained statistically significant, and the direction of effect was consistent, irrespective of which single study was removed. Furthermore, the high level of heterogeneity (I² > 88%) persisted, indicating that no single study disproportionately influenced the overall heterogeneity.

**Figure 3:**
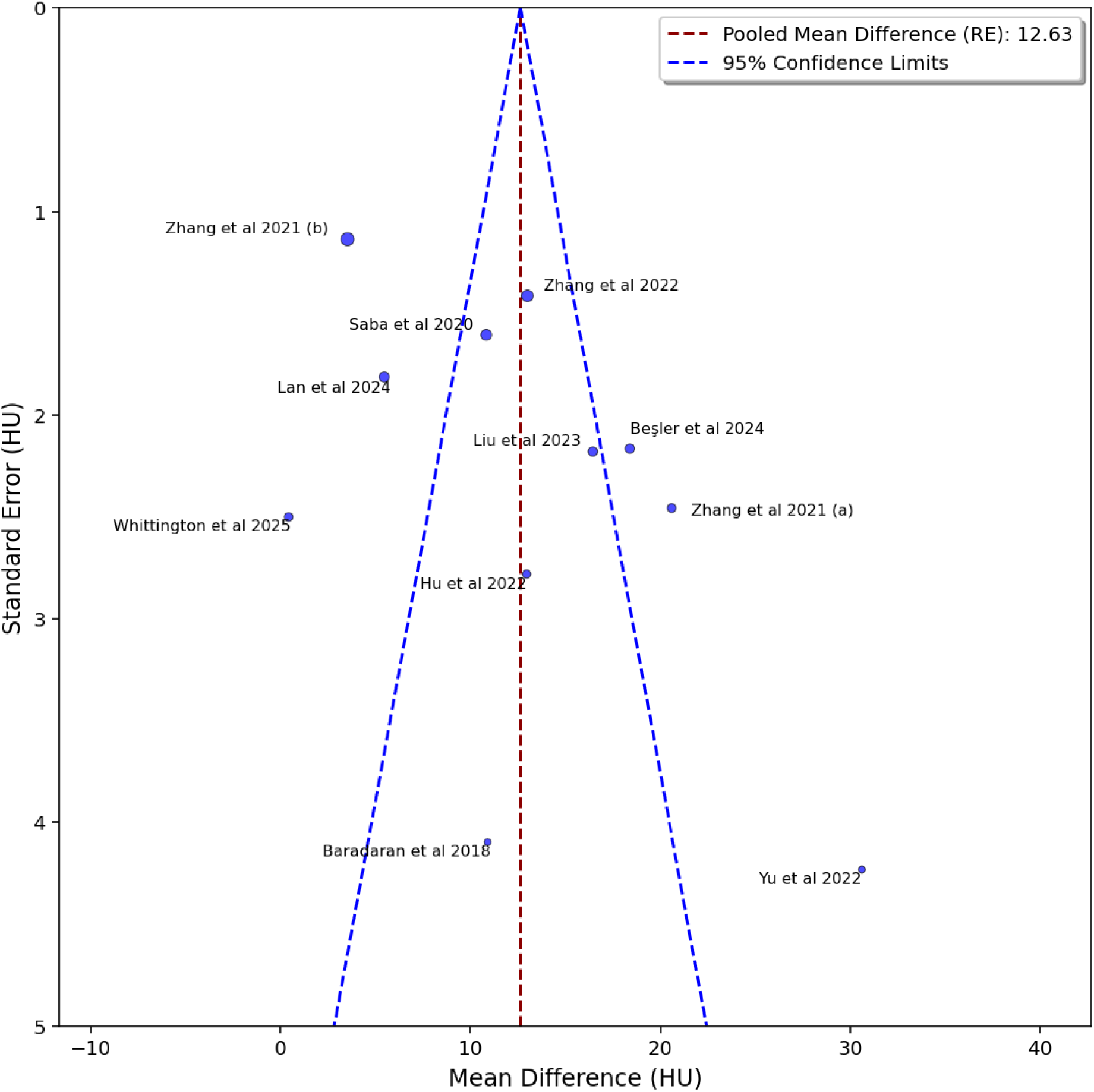
Funnel plot assessing potential publication bias. The x-axis represents the mean difference in perivascular fat attenuation (PVAT) between symptomatic and asymptomatic carotid arteries (Hounsfield Units, HU), and the y-axis represents the standard error (HU). Each circle represents an individual study, labelled by the first author and year of publication. The vertical dashed red line indicates the pooled mean difference from the random-effects model. The blue dashed lines represent pseudo 95% confidence limits around the pooled effect, forming the expected funnel shape in the absence of publication bias.

## Discussion

This meta-analysis, encompassing 11 studies and 2316 patients, assessed whether carotid artery perivascular adipose tissue (PVAT) attenuation on computed tomography angiography (CTA) could serve as an imaging biomarker for symptomatic carotid atherosclerosis. Our pooled analysis demonstrated a statistically significant increase in PVAT attenuation in symptomatic compared to asymptomatic carotid arteries. This mirrors findings in coronary artery disease, where elevated perivascular fat attenuation (often termed fat attenuation index, FAI) links to increased peri-arterial inflammation and adverse cardiovascular outcomes [15]. While carotid-specific evidence directly correlating higher PVAT attenuation with PET-derived inflammation markers or future cerebrovascular events is still limited, our meta-analysis consistently shows strong association with symptomatic carotid arteries. These findings suggest a trend that higher PVAT values correlate with increased plaque instability, which may be used in prospective studies to identify high-risk patients and potentially guide preventative strategies.

The strong link found between higher perivascular fat attenuation and symptomatic carotid arteries reinforces findings from other advanced imaging techniques used to investigate plaque inflammation and instability. For example, fluorodeoxyglucose (FDG) positron emission tomography (PET) shows increased tracer uptake within symptomatic plaques, indicating heightened metabolic inflammatory activity [6, 7]. Similarly, plaque radiomics reveals subtle, quantifiable differences in plaque composition and morphology, pointing to vulnerability.

However, it is important to acknowledge that our overall pooled findings regarding PVAT differ from a recent study by Whittington et al. [17]. This study found no significant difference in carotid PVAT attenuation between culprit and non-culprit internal carotid arteries. The discrepancy between our pooled results and their findings may stem from several factors. Whittington et al. (2025) employed a highly standardised and comprehensive quantitative volumetric approach across all carotid segments. While several studies in our meta-analysis also utilised within-subject controls (comparing symptomatic to asymptomatic sides within the same patient), Whittington et al. (2025) used a particular combination of volumetric analysis and contralateral comparison which may offer different insights compared to the more diverse, predominantly region-of-interest (ROI) based methodologies observed across the other included studies. Among these, 4 out of 11 studies utilised various semi-automated software packages for PVAT calculation, and the remaining relied on manual ROI measurements. The methodological variability, particularly in ROI placement and software used, likely contributes to the differing insights. Furthermore, variations in patient cohort selection and the precise definition of "culprit" or "symptomatic" vessels, as well as differences in sample size and overall study design, likely contribute to these differing results.

Substantial heterogeneity was observed across the included studies (I² = 92%, Q=125.40, p<0.001). The heterogeneity is likely due to a combination of factors, including variations in: (1) patient characteristics (such as severity of carotid stenosis, time since symptom onset, and comorbidities), (2) CT imaging protocols (e.g., scanner settings, reconstruction algorithms), and (3) region of interest (ROI) analysis methodologies (e.g., specific placement of ROI boundaries, inclusion/exclusion of tissues, the use of semi-automatic versus manual measurements, and different analytical software).

Assessment of potential publication bias using a funnel plot revealed some visual asymmetry, raising the possibility of underrepresentation of smaller studies with null or negative findings (Figure 3). Egger’s regression test reinforced concerns regarding potential publication bias by indicating statistically significant asymmetry (intercept = 7.29, p = 0.0440), which could lead to an overestimation of the true effect size of the pooled mean differences. Given the substantial heterogeneity observed, both the visual assessment and Egger’s test should be interpreted with caution. A leave-one-out sensitivity analysis confirmed the stability of our findings with the pooled mean difference remaining statistically significant despite individual study removal.

Several limitations warrant consideration. First, this meta-analysis relies on a relatively small number of observational studies, which naturally limits the real-world generalizability of our findings. Second, our use of aggregated data precluded more granular analyses, such as individual patient risk stratification based on PVAT measurements. Third, it is important to note that all included studies are observational and cross-sectional, assessing PVAT attenuation in patients who have already experienced an ischemic event or are being evaluated for symptomatic disease. Consequently, these studies are descriptive of post-event characteristics rather than prognostic or designed for independent risk stratification for future cerebrovascular events. Fourth, our meta-analysis focused exclusively on mean PVAT attenuation. This approach did not allow for the inclusion or analysis of more sophisticated second-order (or higher-order) quantitative perivascular indices, such as those derived from radiomics, which have shown promise in identifying plaque instability and inflammation in recent studies [19, 20]. Fifth, as evidenced by the high heterogeneity, study design, PVAT analysis methodology, and patient cohort characteristics differed significantly between the included studies. This methodological variability introduces inherent uncertainty into the pooled estimate.

Despite these limitations and the presence of some conflicting signals from individual studies, this meta-analysis provides compelling evidence supporting the potential of PVAT as a non-invasive imaging marker for identifying increased inflammation and vulnerable plaques in symptomatic carotid atherosclerosis. Furthermore, PVAT may serve as a useful biomarker and endpoint to monitor the effects of treatments targeting vascular inflammation in future clinical trials. Further large-scale, prospective studies are needed to: (1) validate the prognostic value of PVAT fat attenuation index for cerebrovascular events, and (2) determine its incremental value in risk stratification beyond established clinical and imaging parameters. These efforts should also aim to establish definitive diagnostic thresholds or cut-off values for PVAT attenuation that could reliably classify carotid plaques as inflamed or vulnerable for clinical decision-making. Ultimately, PVAT attenuation may facilitate the development of personalised prevention strategies for carotid atherosclerosis to reduce the burden of stroke.

## Data Availability

All data produced in the present work are contained in the manuscript.

## Acknowledgments

Nil.

